# Acute tubular necrosis in patients receiving immune check-point inhibitors

**DOI:** 10.64898/2025.12.30.25343098

**Authors:** Juliette Pellegrini, Victor Gueutin, Hassan Izzedine, Lucile Paris, Magali Colombat, Isabelle Brocheriou, Bastien Jean Jacques, Thomas Perier, David Ribes, Antoine Huart, Julie Belliere

**Affiliations:** Department of Nephrology and Organ Transplantation, Referral Centre for Rare Kidney Diseases, French Intensive Care Renal Network, University Hospital of Toulouse, France; Department of Nephrology University Hospital of Caen, France; Nephrology-Dialysis Department, J.Monod Hospital, Flers, France; Department of Nephrology, Peupliers Private Hospital, Ramsay Générale de Santé, Paris, France; Department of Pathology, University Hospital of Toulouse, University Cancer Institute of Toulouse, Toulouse, France; Department of Pathology, Assistance Publique-Hôpitaux de Paris, Hôpital Pitié-Salpétrière, Paris, France; Université Sorbonne, Paris, France; Department of Pathology, University Hospital of Caen, Caen, France; Université Toulouse, Toulouse, France; INSERM U1297, Institute of Metabolic and Cardiovascular Diseases, Toulouse, France

## Abstract

**Background:** Immune checkpoint inhibitor (ICI)–associated acute interstitial nephritis (ICI-AIN) has emerged as a common cause of acute kidney injury (AKI). However, acute tubular necrosis (ATN) may present with similar clinical features yet requires fundamentally different management. Robust data on the prevalence of ATN among patients with suspected ICI-AIN are limited. The aim of this study was to determine the prevalence of ATN in patients with suspected ICI-AIN and to estimate the risk of diagnostic error associated with empirical corticosteroid therapy in the absence of kidney biopsy.

**Methods:** We conducted a retrospective multicenter study including ICI-treated patients who developed AKI and underwent kidney biopsy. Patients with glomerular presentations were excluded. Based on histopathological findings, patients were classified into two groups: ICI-AIN and isolated ATN. The primary outcome was the prevalence of ATN among patients with suspected ICI-AIN. Secondary outcomes included the description of clinical characteristics, concomitant medications, and laboratory findings, as well as the identification of factors potentially distinguishing ATN from ICI-AIN.

**Results:** A total of 132 patients were included. ICI-AIN was diagnosed in 76 patients (58%), while isolated ATN was identified in 56 patients (42%). Among subgroups with known risk factors for ICI-AIN, isolated ATN was observed in 20% (7/35) of patients exposed to proton pump inhibitors (PPIs), 23% (3/13) receiving combination ICI therapy, 31% (5/16) with pre-existing chronic kidney disease (CKD), and 34% (10/34) with concomitant extrarenal immune-related adverse events (irAEs). ATN was also present in 23% (6/26) of patients treated with ICI monotherapy.

Patients with AIN had more severe renal impairment, with a higher median serum creatinine level at presentation (200 µmol/L vs. 141 µmol/L, p < 0.001). Leukocyturia and hematuria were significantly more frequent in the AIN group (p = 0.007 and p = 0.009, respectively). PPI use was significantly more common in patients with AIN than in those with ATN (40% vs. 19%, p = 0.027), whereas exposure to platinum-based chemotherapy was more frequent in the ATN group (78% vs. 59%, p = 0.027).

**Conclusion:** Although certain clinical features are associated with ICI-AIN, they are insufficient to reliably differentiate it from ATN without histological confirmation. An empirical management strategy involving corticosteroid therapy without kidney biopsy carries a diagnostic error risk of approximately 40%, underscoring the importance of kidney biopsy as the diagnostic gold standard.

## Introduction

Immune checkpoint inhibitors (ICI) may induce acute kidney injury (AKI), potentially precluding the use of further chemotherapeutic agents and adversely affecting both renal and overall prognosis. Balancing the priorities of kidney preservation and effective oncologic control remains a major clinical challenge. While ICI-associated acute interstitial nephritis (ICI-AIN) has become a frequent and well-recognized cause of ICI-related AKI^1^, its management warrants particular attention, as alternative diagnoses—most notably acute tubular necrosis (ATN)—may present with similar clinical features.

Because kidney biopsy is invasive and challenging in oncologic patients, particularly in the setting of single kidney, systemic anticoagulation or antiplatelet therapy, clinicians may favor an empirical approach consisting of steroid initiation and ICI discontinuation. However, such a strategy may result in unnecessary steroid exposure, with potential impairment of ICI efficacy, as well as an avoidable interruption of oncologic therapy, thereby compromising cancer outcomes.

To date, robust data on the prevalence of ATN in patients with suspected ICI-AIN are limited^2,3^. In published case series reporting kidney biopsies in ICI-treated patients, the proportion of isolated ATN ranges from 0% to 41.7%^4–6^, with a mean prevalence of approximately 14% (Supplementary Table 1).

We therefore hypothesized that ATN was underestimated and conducted a large multicenter study with the primary objective of determining its prevalence in patients with suspected ICI-AIN, thereby estimating the risk of diagnostic error associated with empirical steroid therapy without kidney biopsy. The secondary objective was to identify factors that might help distinguishing ATN from ICI-AIN patients.

### Short Methods

We retrospectively reviewed the medical records of all patients who received ICI treatment, developed AKI and had a diagnostic kidney biopsy in 3 French centers. Patients were divided into 2 groups (AIN or ATN) according to common diagnostic criteria, respectively presence of a significant immune infiltrate associated with tubulitis, or absence of immune cells and isolated tubular injury features. Full methods are provided in **Supplementary material**.

## Results

### Patient demographics and clinical characteristics

#### Baseline characteristics

We identified a total of 154 patients diagnosed with cancer, treated with ICI, and who developed an AKI requiring a kidney biopsy. Of these, 22 had a glomerular disease and were excluded from the further analyses. Clinical characteristics are provided in **Table 1**. Patients were predominantly males (62.1 %), with a median age of 65 years old. The main comorbidities included hypertension (40%) and diabetes (11%). The most frequent malignancies were lung adenocarcinoma (57%), followed by melanoma (21%) and renal cell carcinoma (5%). The most frequent ICI was an anti-PD1 Pembrolizumab (65.2%), whereas ICI combination represented only 10.6%. Cancer therapy included a platinum-based chemotherapy in 66% of cases. The median number of ICI injections preceding AKI episode was 5 [1-32].

**Table 1.** Characteristics of the global cohort and comparison between ATN vs ICI-AIN patients.

|  | **Total**<br>**n=132<sup>1</sup>** | **ATN**<br>**n=56<sup>1</sup>** | **AIN**<br>**n=76<sup>1</sup>** | **p-value<sup>2</sup>** |
| --- | --- | --- | --- | --- |
| **Baseline parameters** |  |  |  |  |
| **Male gender, n (%)** | 82 (62) | 40 (71) | 42 (55) | 0.058 |
| **Age at time of AKI (y), median (25%,75%)** | 65 (58, 72) | 63.5 (57.5,70) | 65 (59.5,73) | 0.2 |
| **Comorbidities, n (%)** |  |  |  |  |
| Hypertension | 53 (40) | 27 (48) | 26 (34) | 0.10 |
| Diabetes mellitus | 14 (11) | 6 (11) | 8 (11) | > 0.9 |
| Baseline eGFR<60 mL/min/1.73m<sup>2</sup>, n (%) | 16 (12.1) | 5 (8.9) | 11 (14.5) | .42 |
| BMI, median (25%,75%) | 24.5 (21.9, 26.4) | 24 (22.1,27.7) | 24.8 (21.9,26) |  |
| **Malignancy treated with ICI, n (%)** |  |  |  | 0.021 |
| Lung adenocarcinoma | 75 (56.8) | 35 (63) | 40 (53) |  |
| Melanoma | 28 (21.2) | 8 (14) | 20 (26) |  |
| Renal cell carcinoma | 7 (5.3) | 4 (7) | 3 (4) |  |
| Ovarian carcinoma | 3 (2.3) | 0 (0) | 3 (4) |  |
| Others | 19 (14.4) | 9 (16) | 10 (13) |  |
| **ICI type, n (%)** |  |  |  | 0.048 |
| Atezolizumab (anti-PDL1) | 6 (4.5) | 5 (8.9) | 1(1.3) |  |
| Avelumab (anti-PDL1) | 1 (0.8) | 0 (0) | 1 (1.3) |  |
| Cemiplimab (anti-PD1) | 1 (0.8) | 1 (1.8) | 0 (0) |  |
| Dostarlimab (anti-PD1) | 2 (1.5) | 0 (0) | 2 (2.6) |  |
| Durvalumab (anti-PDL1) | 3 (2.3) | 2 (3.6) | 1 (1.3) |  |
| Ipilimumab (anti-CTLA4) | 1(0.7) | 0 (0) | 1 (1.3) |  |
| Nivolumab (anti-PD1) | 31(23.5) | 9 (16) | 22 (29) |  |
| Pembrolizumab (anti-PD1) | 86 (65.2) | 38 (68) | 48 (63) |  |
| ICI used in combination | 14 (10.6) | 3 (5.4) | 11 (14) | 0.093 |
| **Comedications, n (%)\*** |  |  |  |  |
| PPI | 35/107 (32.7) | 7/37 (19) | 28/70 (40) | 0.027 |
| NSAID | 4/107 (3.7) | 2/37 (5.4) | 2/70 (2.9) | 0.6 |
| Antibiotics | 15/107 (14) | 1/37 (2.7) | 14/70 (20) | 0.014 |
| Iodine contrast products | 7/105 (66.7) | 0/35 (0) | 7/70 (10) | 0.092 |
| Platinum-based chemotherapy | 87 (66) | 42 (78) | 45 (59) | 0.027 |
| Pemetrexed only | 68 (52) | 31 (55.4) | 37 (48.6) | 0.48 |
| Chemotherapies with platinum+pemetrexed | 88 (67) | 42 (75) | 46 (60.5) | 0.094 |
| **At admission for AKI** |  |  |  |  |
| Number of injections before AKI, median (25%,75%) | 4.5 (3,9.3) | 4.5 (3,7.5) | 4.5 (3,9.8) | 0.7 |
| Time between first inj. and AKI (weeks), median (25%,75%) | 18 (9,32.8) | 19.0 (9.8,26.8) | 17.5 (9,33.8) | > 0.9 |
| Time between last inj. and AKI (weeks), median (25%,75%) | 2 (1,3) | 2 (1,3) | 3 (1,3) | 0.2 |
| extra-renal IRAe, n (%)* | 34/98 (34.7) | 10/28 (36) | 24/70 (34) | 0.9 |
| **Biological parameters** |  |  |  |  |
| SCr creatinine (μmol/L), median (25%,75%) | 162 (136, 242) | 141 (129,173) | 200 (153,300) | < 0.001 |
| Proteinuria (g/g), median (25%,75%) | 0.3 (0.1,0.4) | 0.2 (0.1,0.3) | 0.3 (0.2,0.4) | 0.062 |
| Hematuria (/mm<sup>3</sup>), n (%)* | 40/117 (34.2) | 9/46 (20) | 31/71 (44) | 0.007 |
| Leucocyturia (/mm<sup>3</sup>), n (%)* | 57/115 (50) | 15/44 (34) | 42/71 (59) | 0.009 |
| Hypereosinophilia, n (%)* | 10/93 (10.8) | 3/31 (9.7) | 7/62 (11) | > 0.9 |
| CRP (mg/L), median (25%,75%) | 10.2 (3.4, 58) | 6 (3.4,11.3) | 20 (4,66) | 0.052 |

<sup>1</sup>n/N (%) <sup>2</sup>Wilcoxon rank sum test; Fisher's exact test; Pearson's Chi-squared test
\* indicate missing data. All other data are complete.
AKI, acute kidney injury; CTLA-4, cytotoxic T lymphocyte-associated antigen 4; eGFR, estimated glomerular filtration rate; ICI, immune checkpoint inhibitor; irAEs, immune-related adverse events; NSAID, non-steroidal anti-inflammatory drug; PD-1, programmed cell death 1; PD-L1, programmed death-ligand 1; PPI, proton pump inhibitor; SCr, serum creatinine.

### AKI presentation

Only 12% of patients were known to have chronic kidney disease (CKD) defined with estimated glomerular filtration rate (eGFR) < 60 mL/min/1.73m^2^. At admission, AKI was severe with a median serum creatinine reaching 162 µmol/L. Proteinuria was low, with only 20% of patients presenting with protein creatinine ratio > 0.5g/g. Leukocyturia and hematuria were reported in 50% and in 34% of cases, respectively.

### ATN prevalence and known risk factors of ICI-AIN

According to biopsy-proven diagnosis, patients were split into 2 groups, corresponding either to ICI-AIN or ATN. As depicted in **Figure 1**, in our global cohort (n=132), ICI-AIN was identified in 76 patients (58%) and ATN in 56 patients (42%). In subgroups of patients with a known risk factor for ICI-AIN^5,6^, isolated ATN occurred in 20% (7/35) of patients with PPI exposure, 23% (3/13) receiving combination ICI therapy, 31% (5/16) with pre-existing (CKD), and 34% (10/34) with presence of extrarenal immune-related adverse events (irAEs). ATN was also observed in 23% (6/26) of patients treated with ICI monotherapy.

**Figure 1.**
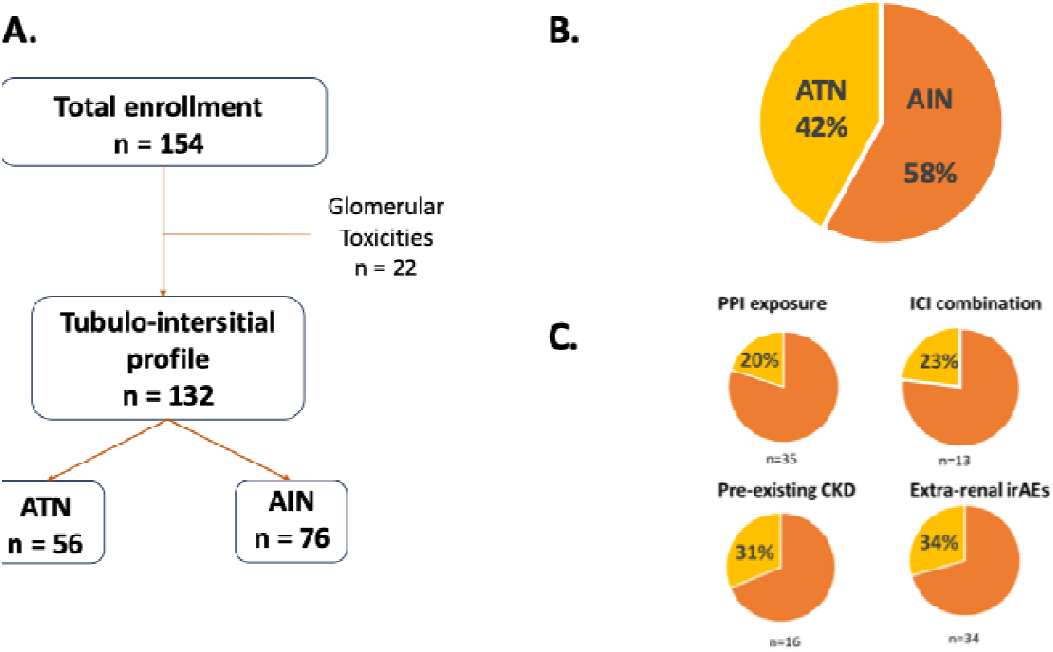
Acute tubular necrosis incidence in patients receiving immune check-point inhibitors (ICI). **Panel A**. Flow-chart of the study. ATN, acute tubular necrosis, AIN, acute interstitial nephritis. **Panel B**. Proportion of ATN in AKI patients with biopsy-proven diagnosis in the whole cohort. **Panel C**. Proportion of ATN patients in sub-group of patients with known risk factor of AIN. PPI, proton-pump inhibitors; CKD, chronic kidney disease; irAEs, extra-renal immune related adverse events.

### Identification of discriminant factors between ATN and AIN patients

We then aimed to determine whether clinical or biological features could differentiate ICI-AIN from ATN patients (**Table 1**).

### Clinical characteristics prior to AKI

Regarding cancer type, lung cancer was more prevalent in the ATN group (62% vs. 53%, p = 0.021). The distribution of ICI regimens (as well as ICI-combination) was similar between groups. Exposure to platinum-based chemotherapy was higher in ATN group compared to AIN group (78% vs 59%, p=0.027). No difference was noted regarding exposure to pemetrexed (50%). The proportion of patients receiving PPI was significantly higher in the AIN group than in the ATN group (40% vs. 19%, p = 0.027). Exposure to antibiotics and iodinated contrast agents was also more frequent among patients with AIN compared with those with ATN (p=0.014 and 0.092, respectively).

### Clinical characteristics at AKI occurrence

The presence of extrarenal irAEs was not distinguishing ATN from ICI-AIN patients (36% vs 34%, p=0.9). Neither the number of ICI injections, nor the time from initiation of ICI or from the last infusion differed significantly. However, patients with AIN presented with more severe renal impairment, as reflected by a higher median serum creatinine level at admission (200µmol/L vs. 141µmol/L, p< 0.001). Leukocyturia and hematuria were significantly more frequent in patients with AIN, with p-values reaching 0.007 and 0.009, respectively, whereas hypereosinophilia, proteinuria, C-reactive protein did not discriminate between groups.

## Discussion

The aim of our study was to assess the prevalence of isolated ATN in patients with ICI-AKI, in order to quantify the risk of diagnostic misclassification when empirical corticosteroid therapy is initiated without kidney biopsy.

In this large multicenter study relying only on biopsy-proven cases, isolated ATN accounted for 42% of diagnoses. Notably, ATN prevalence was meaningful (20-32%) in sub-groups of patients with well-known AIN risk factors (PPI exposure, ICI combination, previous CKD, extra-renal irAEs), as well as in patients exposed to ICI in monotherapy (without nephrotoxic chemotherapy).

Our study population was very close to previously published cohorts regarding oncological characteristics and clinical presentation^6,7^. Patients with ICI-AIN presented with more severe renal involvement, were more likely to exhibit leukocyturia and hematuria, and had more frequently been exposed to potential confounding medications, including PPI, antibiotics, and iodinated contrast agents. In contrast, patients with ATN were more often exposed to nephrotoxic chemotherapy and exhibited less severe renal dysfunction. Our real-world data demonstrate a higher prevalence of ATN than previously reported^8^. This could be explained by a larger use of ICI and a better knowledge of ICI-AKI, prompting patients to be referred to nephrology units, and the recent recommendations promoting kidney biopsy^8^.

The retrospective design entails inherent limitations, including missing data preventing us to perform a multivariate analysis. An indication bias for kidney biopsy cannot be ruled out as well, and the absence of centralized pathological review may have also introduced interobserver variability. Nevertheless, the implications of this study are substantial: our data suggest that empiric strategy in ICI-AKI management leads to an overtreatment and an irrelevant discontinuation of ICI in more than 1 patient out of 3, advocating for a systematic kidney biopsy, when feasible, in every ICI-AIN suspicion to obtain a clear diagnosis.

While suspecting ICI-AIN, clinical criteria alone appear insufficient to avoid kidney biopsy (the current gold standard). In the absence of routinely available ICI-AIN urinary biomarkers^9^, both clinicians and patients deserve to be aware that an empiric strategy including steroids prescription without histological features carries a 40% error risk.

## Data Availability

All data produced in the present work are contained in the manuscript

## Supplementary Material section

Supplementary File (PDF) includes:

- Supplementary Methods
- Supplementary Table 1 reporting the reported prevalence of ATN in case of AIN suspicion
- Supplementary references

## References

1. Sprangers B, Leaf DE, Porta C, Soler MJ, Perazella MA. Diagnosis and management of immune checkpoint inhibitor-associated acute kidney injury. Nat Rev Nephrol. 2022;18(12):794–805. doi:10.1038/S41581-022-00630-8

2. Cortazar FB, Marrone KA, Troxell ML, Ralto KM, Hoenig MP, Brahmer JR, L. DT, Lipson EJ, Glezerman IG, Wolchok J, Cornell LD, Feldman P, Stokes MB, Zapata SA, Hodi FS, Ott PA, Yamashita M LDE. Clinicopathological features of acute kidney injury associated with immune checkpoint inhibitors. - PubMed - NCBI. Kidney Int. 2016;S0085-2538(16):30164–30168. doi:10.1016/j.kint.2016.04.008

3. Cortazar FB, Kibbelaar ZA, Glezerman IG, et al. Clinical Features and Outcomes of Immune Checkpoint Inhibitor-Associated AKI: A Multicenter Study. J Am Soc Nephrol. 2020;31(2):435–446. doi:10.1681/ASN.2019070676

4. Izzedine H, Mathian A, Champiat S, et al. Renal toxicities associated with pembrolizumab. Clin Kidney J. 2019;12(1):81–88. doi:10.1093/ckj/sfy100

5. Cortazar FB, Kibbelaar ZA, Glezerman IG, et al. Clinical Features and Outcomes of Immune Checkpoint Inhibitor-Associated AKI: A Multicenter Study. J Am Soc Nephrol. 2020;31(2):435–446. doi:10.1681/ASN.2019070676

6. Gupta S, Short SAP, Sise ME, et al. Acute kidney injury in patients treated with immune checkpoint inhibitors. J Immunother Cancer. 2021;9(10). doi:10.1136/JITC-2021-003467

7. Cortazar FB, Marrone KA, Troxell ML, et al. Clinicopathological features of acute kidney injury associated with immune checkpoint inhibitors. Kidney Int. 2016;90(3):638–647. doi:10.1016/j.kint.2016.04.008

8. Herrmann SM, Abudayyeh A, Gupta S, et al. Diagnosis and management of immune checkpoint inhibitor-associated nephrotoxicity: a position statement from the American Society of Onco-nephrology. Kidney Int. 2025;107(1):21–32. doi:10.1016/J.KINT.2024.09.017

9. Long JP, Singh S, Dong Y, Yee C, Lin JS. Urine proteomics defines an immune checkpoint-associated nephritis signature. J Immunother Cancer. 2025;13(1). doi:10.1136/jitc-2024-010680

