## Supplementary material for "Acute tubular necrosis in patients receiving immune check-point inhibitors"

**Full methods section**

***Patients***

We retrospectively selected patients exposed to ICI, who were referred for kidney biopsy to nephrology units (Toulouse, Caen, Paris) between 2015 and 2025. Clinical and biological data were collected from medical records and included age, gender, type of malignancy, comorbidities, current treatment (ICI and comedications). At admission for AKI, we reviewed for each patient: creatinine, estimated glomerular filtration rate (eGFR) calculated with CKD-EPI formula, and urinalysis.

***Ethics***

The study was conducted according to MR-004 rule adopted by the French data protection authority ("CNIL") for the processing of personal data within the context of an observational study and declared under the reference RC31/25/0116 at Toulouse University Hospital and 3155bis/2022 for Caen/Paris.

***Definitions***

AKI was diagnosed based on common terminology criteria KDIGO stages. AIN and ATN were defined with common histopathological criteria[9]: (i) presence of mononuclear cells in the basolateral aspect of the renal tubule epithelium and/or tubulitis for AIN (infiltration of lymphocytes, macrophages, and eosinophils in the renal interstitium); (ii) isolated tubular epithelial flattening, tubular dilatation, tubular cell necrosis, regenerative changes, tubular cell vacuolization, tubular cell sloughing and loss of the brush border for ATN. The absence of significant immune infiltration was the required criteria to diagnose ATN.

***Statistical analysis***

Statistical analyses were performed using R software (version 4.2.0). Patient characteristics were summarized using descriptive statistics, including frequency distributions and medians (range). Group comparisons were conducted using Chi-square or Fisher's exact tests for categorical variables, and the Wilcoxon test for continuous variables. A p-value $< 0.05$ was considered statistically significant. Given the proportion of missing data for some key variables and the instability observed in the multivariable models (highly variable coefficients, wide confidence intervals, and quasi-separation issues), we decided not to retain the multivariable analysis. A comprehensive univariable analysis was preferred, as it was deemed more robust given the quality of the dataset.

**Supplementary references**

S1. Izzedine H, Mathian A, Champiat S, et al. Renal toxicities associated with pembrolizumab. Clin Kidney J. 2019;12(1):81-88. doi:10.1093/ckj/sfy100

S2. Cassol C, Satoskar A, Lozanski G, et al. Anti−PD-1 Immunotherapy May Induce Interstitial Nephritis With Increased Tubular Epithelial Expression of PD-L1. Kidney Int Rep. 2019;4(8):1152-1160. doi:10.1016/j.ekir.2019.06.001

S3. Mamlouk O, Selamet U, Machado S, et al. Nephrotoxicity of immune checkpoint inhibitors beyond tubulointerstitial nephritis: single-center experience. J Immunother Cancer. 2019;7(1):2. doi:10.1186/s40425-018-0478-8

S4. Gérard AO, Andreani M, Fresse A, et al. Immune checkpoint inhibitors-induced nephropathy: a French national survey. Cancer Immunol Immunother. 2021;70(11):3357-3364. doi:10.1007/S00262-021-02983-8

S5. Singh S, Long JP, Tchakarov A, Dong Y, Yee C, Lin JS. Tertiary lymphoid structure signatures are associated with immune checkpoint inhibitor related acute interstitial nephritis. JCI Insight. Published online December 1, 2022. doi:10.1172/JCI.INSIGHT.165108

S6. Jin S, Shen Z, Li J, et al. Clinicopathological features of kidney injury in patients receiving immune checkpoint inhibitors (ICPi) combined with anti-vascular endothelial growth factor (anti-VEGF) therapy. J Clin Pathol. 2024;77(7):471-477. doi:10.1136/JCP-2023-209173

S7. Kommer A, Stortz M, Kraus D, Weinmann-Menke J. Immune Checkpoint Inhibitor-Associated Acute Kidney Injury: A Single-Center Experience of Biopsy-Proven Cases. J Clin Med. 2025;14(9). doi:10.3390/JCM14093231
S8. Al-Othman YA, Metcalf BD, Kroneman O, et al. Grading T Lymphocyte-Mediated Acute Interstitial Nephritis Following Checkpoint Inhibitor Therapy. Ann Clin Lab Sci. 2025;55(2):172-178. Accessed December 24, 2025. https://pubmed.ncbi.nlm.nih.gov/40340877/
S9. Sprangers B, Leaf DE, Porta C, Soler MJ, Perazella MA. Diagnosis and management of immune checkpoint inhibitor-associated acute kidney injury. Nat Rev Nephrol. 2022;18(12). doi:10.1038/S41581-022-00630-8

**Supplementary Table 1. ATN prevalence in previously reported kidney biopsies from patients with ICI-related AKI.**

| Reference | Reported Kidney Biopsies | ATN | AIN | Other diagnosis | ATN prevalence |
|---|---|---|---|---|---|
| Cortazar *et al.* 2016 [2] | 13 | 0 | 12 | 1 | 0 |
| Izzedine *et al.* 2019[S1] | 12 | 5 | 4 | 3 | 41,7 |
| *Cassol et al. KIR* 2019[S2] | 15 | 5 | 9 | 1 | 33 |
| *Mamlouk et al.* 2019[S3] | 16 | 1 | 14 | 1 | 6 |
| *Cortazar et al.* 2020[3] | 60 | 0 | 56 | 4 | 0 |
| *Gérard et al.* 2021[S4] | 63 | 6 | 52 | 5 | 10 |
| Gupta *et al.* 2021[6] | 151 | Figure 2F | 125 | NA | < 8 |
| *Singh et al.* 2022[S5] | 36 | 9 | 18 | 9 | 25 |
| *Jin et al.* 2024[S6] | 12 | 0 | 9 | 3 | 0 |
| *Kommer et al. 2025*[S7] | 12 | 0 | 12 | 0 | 0 |
| *Al-Othman et al. 2025*[S8] | 20 | 5 | 14 | 1 | 26 |
